# Self-reported symptoms of covid-19 including symptoms most predictive of SARS-CoV-2 infection, are heritable

**DOI:** 10.1101/2020.04.22.20072124

**Authors:** Frances MK Williams, Maxim B Freidin, Massimo Mangino, Simon Couvreur, Alessia Visconti, Ruth CE Bowyer, Caroline I Le Roy, Mario Falchi, Carole Sudre, Richard Davies, Christopher Hammond, Cristina Menni, Claire J Steves, Tim D Spector

## Abstract

Susceptibility to infection such as SARS-CoV-2 may be influenced by host genotype. TwinsUK volunteers (n=2633) completing the C-19 Covid symptom tracker app allowed classical twin studies of covid-19 symptoms including predicted covid-19, a symptom-based algorithm predicting true infection derived in app users tested for SARS-CoV-2. We found heritability for fever = 41 (95% confidence intervals 12-70)%; anosmia 47 (27-67)%; delirium 49 (24-75)%; and predicted covid-19 gave heritability = 50 (29-70)%.

## Introduction

Infectious diseases may demonstrate a heritable component – that is the propensity to contract and develop active infection and the severity of the immune response - is influenced by host genetic factors. Viral diversity is associated with genetic variants mediating the immune response and biosynthesis of glycan structures functioning as virus^1^ and immunogenetic factors are implicated in risk and severity of H1N1 infection^2^. Understanding how symptoms of covid-19 pass through the population may indicate the pathogenic mechanisms of SARS-CoV-2 infection^3^ as well as offering utility in the allocation of scarce healthcare resources, particularly intensive care beds^4^. We developed the C-19 Covid Symptom Tracker app (https://doi.org/10.1101/2020.04.02.20051334) to collect real time data during the SARS-CoV-2 pandemic. Ethics committee approval was obtained for TwinsUK from St Thomas’ Hospital Ethics Committee 2008 with further approval obtained to use health records for research. The aim of the current study was to estimate the heritability of covid-19 symptoms and identify phenotypes suitable for genome-wide association study.

## Methods

The app asks on a daily basis about the presence or absence of common symptoms including cough, fever, chest pain, delirium, and anosmia and has downloaded by >2 million people. Participants in the TwinsUK adult twin register^5^ who had reported current symptoms via the app were included. Heritability of individual symptoms and ‘Predicted covid-19’ was estimated using the liability threshold model^6^ implemented in the *mets* package for R^7^. The liability-threshold model assumes an underlying continuous liability that follows a normal distribution. The model decomposes observed phenotypic variance into three latent sources of variation: additive genetic (A), common environment (C), and unique environment variance (E) and univariate model comparisons were conducted using χ2 tests. Heritability (A) was estimated from the most parsimonious model (with adjustment for age, sex, and body mass index) as determined by the Akaike information criterion^8^. ‘Predicted covid-19’ status was determined by linear combination of self-reported age, sex and symptoms of anosmia, severe or significant persistent cough, fatigue and skipped meals. We have shown (n= 5238 app users) this model to be most predictive of SARS-CoV-2 positivity following a swab PCR-test (https://doi.org/10.1101/2020.04.05.20048421). We also examined real time information obtained from TwinsUK participants by SMS text regarding their current co-habiting arrangement (n=2,449). To control for the influence of infection within households on estimates of C, we repeated the heritability estimates excluding twins living together (total n = 345; MZ n = 277; DZ n = 68) and those with co-habiting information missing (n = 184).

## Results

Adult same-sex twins (n=2,633) had provided data for analysis between 25^th^ March –April 3^rd^ 2020. Repeat measures have been summarised across all logging occasions: an individual reporting a symptom at any time point was considered a case, otherwise they were allocated control status for that symptom. The sample comprised 728 complete pairs of which 537 pairs were monozygotic and 191 pairs dizygotic, with 86.9% being female. Prevalence of symptoms in TwinsUK is shown in Figure 1 and was similar to the larger dataset of n=1.85 million (data not shown).

**figure 1.**
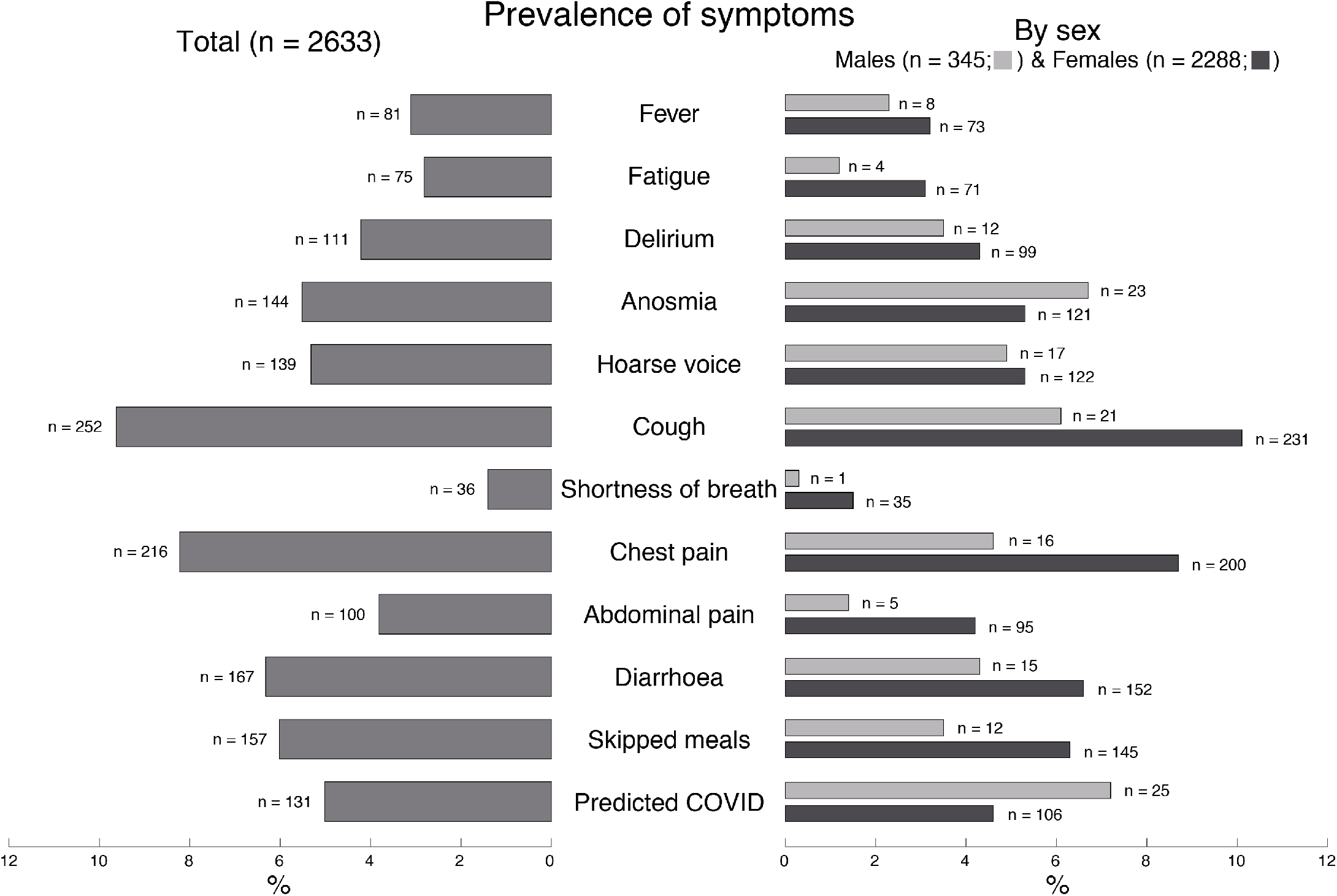
Prevalence of symptoms “Predicted covid-19” status reported by 2633 twins. MZ represents monozygotic; DZ dizygotic twins.

Figure 2 shows the heritability determined by fitting liability threshold models containing additive genetic (A), shared environment (C) and unique environmental (E) factors. A heritable component was seen for delirium (49%, 95% confidence interval = [24-75]%), fever (41%, [12-70]%), fatigue (32 [1-64]%), anosmia (47 [27-67]%), shortness of breath (43 [8-77]%), and diarrhoea (34 [14-55]%) but not for hoarse voice, cough, skipped meals, chest pain, and abdominal pain which were environmentally determined. Heritability estimates including only those twin pairs living apart (n = 2,104) were largely similar, with fever (41 [7-74]%), anosmia (48 [25-72]%), and shortness of breath (47 [12-82]%), increased for diarrhoea (41 [18-63]%), but reduced for delirium (37 [5-68]%) and fatigue (24 [0-61]%). A heritable component was also observed for ‘predicted covid-19’: 50 [29-70]% in the overall sample and 54 [30-77]% in the sample excluding cohabiting and twins missing cohabiting data.

**figure 2.**
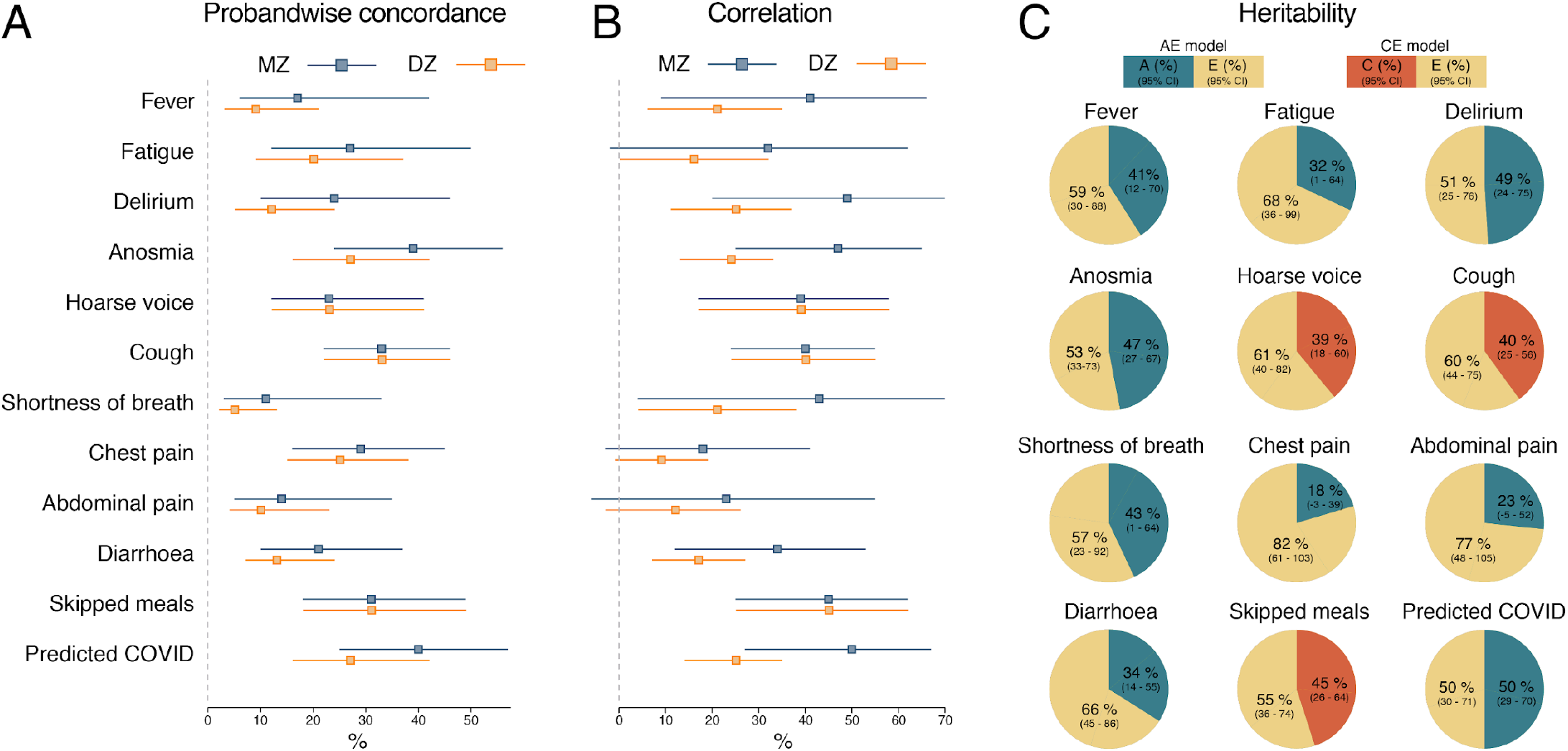
Estimates of probandwise concordance (A), tetrachoric correlation (B), and heritability (C) for 2,633 twins. MZ represents monozygotic; DZ dizygotic; CI confidence interval; A, additive genetic; C, shared environmental; E, unique environment.

## Discussion

Here we report that 50% of the variance of ‘predicted covid-19’ phenotype is due to genetic factors. The current prevalence of ‘predicted covid-19’ is 2.9% of the population. Symptoms related to immune activation such as fever, delirium and fatigue have a heritability >35%. The symptom of anosmia, that we previously reported to be an important predictive symptom of covid-19, was also heritable at 48%. Symptomatic infection with SARS-CoV-2, rather than representing a purely stochastic event, is under host genetic influence to some extent and may reflect inter-individual variation in the host immune response. Viral infections typically lead to T cell activation with IL-1, IL-6 and TNF-α release causing flu-like symptoms such as fever. The genetic basis of this variability in response will provide important clues for therapeutics and lead to identification of groups at high risk of death, which is associated with a cytokine storm at 1-2 weeks after symptom onset^9^.

Our study has several strengths. TwinsUK and their symptom reporting is representative of the UK population. Predicting infection with a validated combination of symptoms based on a large training set (n=5,238) of virus-tested individuals offers a pragmatic solution to the challenges of widespread testing in the general population. The limitations include first, there is likely to be healthy volunteer bias. That said, our study was well powered to find moderate heritability. Second, many symptoms are non-specific and are prevalent in spring in the Northern hemisphere when allergies and seasonal flu are active, and are not indicative of infection status. We did have sufficient sample size of people reporting symptoms and viral test results to indicate accurately which symptom pattern provides the greatest positive predictive value. Our results could have been biased by MZ twins cohabiting more than DZ twins, but real time data collection allowed us to exclude cohabiting pairs. Finally, our twin sample is predominantly female for historical recruitment reasons and is not representative of non-European ancestries ^5^.

The genetic influence on covid-19 symptoms may reflect genotype status of candidate genes such as *ACE2R* which encodes the target for viral attachment^10^. Further genetic work is underway to determine whether twins’ genotype at *ACE2R* influences either predicted positivity or symptoms and a global genetic study is underway (https://www.covid19hg.org/). Public health measures to identify those at increased genetic risk of severe infection would be useful as a way of mitigating the economic effects of lockdown and social distancing policies.

## Data Availability

The data are available to bona fide researchers on request to TwinsUK at King's College London.

## Acknowledgements

TwinsUK is funded by the Wellcome Trust, Medical Research Council, European Union, Chronic Disease Research Foundation (CDRF), Zoe Global Ltd and the National Institute for Health Research (NIHR)-funded BioResource, Clinical Research Facility and Biomedical Research Centre based at Guy’s and St Thomas’ NHS Foundation Trust in partnership with King’s College London. We are grateful to all the users of C-19 Covid Symptom Tracker app for their contribution to this study.

## Author contributions

FMKW, CJS and TDS conceived the study; FMKW produced the first draft of the manuscript; MBF, MM, and SC carried out statistical analysis; AV, RCEB, CILR, MF, CS, RD, CH and CM provided data, computational support and contributed to the results interpretation. All authors read and approved the final version of the manuscript.

## Notes

### Competing Interest Statement

TDS is consultant to Zoe Global Ltd ("Zoe"). RD is an employee of Zoe Global Limited. Other authors have no conflict of interest to declare.

